# Unveiling the pathogenesis of non-alcoholic fatty liver disease by decoding biomarkers through integrated single-cell and single-nucleus profiles

**DOI:** 10.1101/2023.10.05.23296635

**Authors:** Wenfeng Ma, Xin Zhong, Benqiang Cai, Mumin Shao, Xuewen Yu, Minling Lv, Shaomin Xu, Bolin Zhan, Qun Li, Mengqing Ma, Mikkel Brejnholt Kjær, Jinrong Huang, Yonglun Luo, Henning Grønbæk, Lin Lin

## Abstract

**Background and Aims:** Non-alcoholic fatty liver disease (NAFLD) is a heterogenous liver disease encompassing pathological changes ranging from simple steatosis, inflammation and fibrosis to cirrhosis. To further unravel NAFLD pathogenesis, we aimed to decode the candidate NAFLD biomarkers associated with NAFLD severity using publicly available single-cell RNA sequencing (scRNA-seq) and single-nucleus RNA sequencing (snRNA-seq) data.

**Methods:** Seurat v5 and anchor-based reciprocal principal components analysis (RPCA) integration were performed to integrate and analyze the scRNA-seq and snRNA-seq data of 82 liver and Peripheral Blood Mononuclear Cell (PBMC) specimens from NAFLD patients and healthy controls to decode the candidate NAFLD biomarkers generated previously. Using the ‘CellChat’ R package, we analyzed ligand-receptor interactions of our candidate biomarkers from secreted genes to understand their signaling crosstalk and implications in NAFLD’s biological processes.

**Results:** We generated a database (https://dreamapp.biomed.au.dk/NAFLD-scRNA-seq/) to present the NAFLD pathogenesis by analyzing integrated scRNA-seq and snRNA-seq data. Through cell-level decoding, we discovered the expression distribution of the candidate biomarkers associated with NAFLD severity. The analysis of ligand-receptor pairs in NAFLD liver and PBMC data suggests that the IL1B-(IL1R1+IL1RAP) interaction between liver monocytes and hepatocytes/cholangiocytes may explain the correlation between NAFLD severity and IL1RAP down-regulation.

**Conclusions:** We confirmed a strong correlation between liver QSOX1/IL1RAP concentrations and NAFLD severity at the cellular level. Additionally, our analysis of comprehensive data unveiled new aspects of NAFLD pathogenesis and intercellular communication through the use of scRNA and snRNA sequencing data. (ChiCTR2300073940).

**Highlights:**

- Integrated single-cell and single-nucleus profiles from 82 liver and PBMC specimens comprising NAFLD patients and healthy controls with increasing severity were utilized to unveil the NAFLD pathogenesis through decoding candidate biomarkers of NAFLD.
- In cell-level observations, we decoded 16 up-regulated and 22 down-regulated secreting genes previously identified as associated with increasing NAFLD severity in the liver RNA-seq and plasma proteomics data.
- QSOX1, enriched in fibroblasts, and IL1RAP, enriched in hepatocytes, have been further validated and interpreted in integrated single-cell and single-nucleus profiles for their potential to predict NAFLD severity.
- The analysis of intercellular crosstalk, focusing on secreted signaling from our previously identified candidate biomarkers sourced from secreted genes, highlighted the IL1B-(IL1R1+IL1RAP) pathway between liver monocytes and hepatocytes/cholangiocytes. This suggests that this pathway might be a potential reason for the observed downregulation of IL1RAP in NAFLD liver.

**Lay Summary:** We integrated single-cell RNA sequencing (scRNA-seq) and single-nucleus RNA sequencing (snRNA-seq) data to unravel non-alcoholic fatty liver disease (NAFLD) pathogenesis. We decoded candidate biomarkers associated with NAFLD progression, which were previously screened from RNA sequencing (RNA-seq) data of 625 liver samples with a novel gene clustering method. A new version of the R package ‘’Seurat v5’ and anchor-based reciprocal principal components analysis (RPCA) integration were performed to process and integrate scRNA-seq and snRNA-seq data of 82 liver and Peripheral Blood Mononuclear Cell (PBMC) specimens from NAFLD patients and healthy controls. The research delved deeper into the cellular expression patterns of the candidate biomarkers and examined the intercellular communication of their secreted signaling.

**Graphical abstract:** 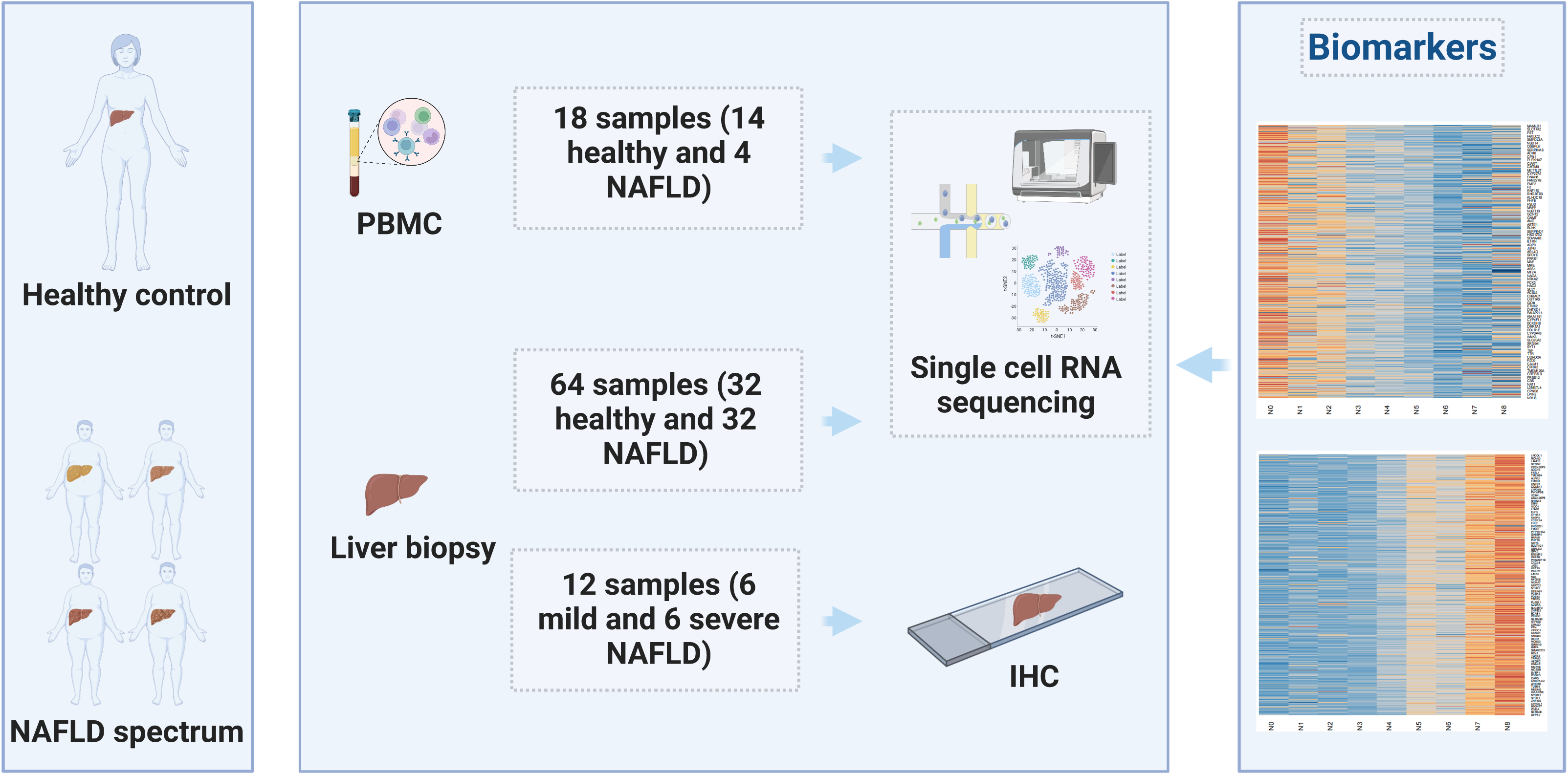

## Introduction

Non-alcoholic fatty liver disease (NAFLD) is a multifaceted disease representing a spectrum from Non-alcoholic Fatty Liver (NAFL) of simple steatosis, non-alcoholic steatohepatitis (NASH) to NASH-induced fibrosis and cirrhosis (1, 2). As the global prevalence of NAFLD reached 38.0%, the upward trend in NAFLD and NASH-related liver cancer balanced the declining tendency of virally-caused hepatocellular carcinoma (3–5). Despite the noteworthy advancements in NAFLD research, the lack of FDA-approved treatments underscores the need for a systematic study to understand its pathogenesis (6, 7).

The advent of transcriptomics, specifically single-cell RNA sequencing (scRNA-seq) and single-nucleus RNA sequencing (snRNA-seq) has triggered a paradigm shift in our comprehension of biological processes, cellular diversity, and the intricacies of physiological and disease states (8). These groundbreaking technologies allow us to examine gene expression at the cell level, significantly enhancing our understanding of various aspects of life sciences (9, 10). In 2019, Xiong et al. applied scRNA-seq to reveal the cellular heterogeneity of NAFLD, identifying key cell types and crosstalk during disease progression (11). Tamburini et al. focused on liver lymphatic endothelial cells (LECs) and observed the activation of the IL13 pathway in LECs of NASH patients (12). In 2020, Ramachandran et al. utilizing single-cell transcriptomics, resolved human liver tissue and blood samples of patients with cirrhosis and healthy controls, identifying new pathogenic subpopulations and providing the markers of cell subtypes (13). In 2021, Deczkowska and colleagues discovered an abundance and activation of type 1 conventional dendritic cells (cDC1) in both NAFLD and NASH conditions with scRNA-seq. They further highlighted XCR1+ cDC1 as a significant contributor to liver pathology in NASH (14). Recently, Wang et al. interpreted NASH-associated hepatic stellate cells (HSCs) with snRNA-seq and uncovered an autocrine HSC signaling circuit that emerged in advanced fibrosis (15). These studies underscore the immense potential of transcriptomics application in NAFLD research for finding molecular signatures and pathways that could guide the development of diagnosis and therapeutics for NAFLD.

Recently, we demonstrated that Quiescin sulfhydryl oxidase 1 (QSOX1) and Interleukin-1 receptor accessory protein (IL1RAP) hold potential as indicators of liver disease severity in NAFLD patients (16). QSOX1, a flavin-linked enzyme that facilitates disulfide bond formation within the secretory structures of mammalian cells, is expelled into the extracellular matrix (ECM) and blood as an independent protein through the standard secretory pathway, encompassing the Golgi apparatus, secretory vesicles, and the plasma membrane. Furthermore, it is notably amplified and released from fibroblasts that are actively synthesizing ECM precursors (15–17). On the other hand, IL1RAP, a member of the IL-1 receptor family, is ubiquitously present but shows high levels in the liver, placenta, and white blood cells, according to databases such as NCBI. This protein plays a critical role in inflammation and is implicated in the initiation and progression of cancer. Studies suggested that both QSOX1 and IL1RAP hold promise as potential biomarkers of NAFLD or obesity (17, 18).

In 2022, Hao’s team unveiled Seurat v5, featuring a ‘bridge integration’ technique designed to merge single-cell datasets from diverse molecular domains and various measurement modalities. Employing concepts from ‘dictionary learning’ within the field of representation learning, this method is highly scalable and ideal for handling expansive datasets of millions of cells. It has wide applicability across different technologies and molecular modalities, making it valuable for large research consortia with existing single-cell RNA-seq databases. Additionally, the approach shows potential in the identification and comprehensive study of rare cellular populations. Overall, the introduction of bridge integration in Seurat v5 is poised to make a significant impact on the advancement of single-cell research (19). Compared to its predecessor (20), Seurat v5 enhanced its capacity for large-scale data integration and facilitated a more comprehensive analysis of the cellular and nuclear transcriptomic landscapes. With Seurat v5, we aimed to use publicly available scRNA-seq and snRNA-seq data to decode the candidate NAFLD biomarkers we have previously screened for unraveling NAFLD pathogenesis.

Understanding global cellular interactions necessitates precise representation and analysis of cell-to-cell signaling pathways. “CellChat” serves as a tool, crafted to extract and evaluate intercellular communication networks from sc- and snRNA-seq data (21). It identifies primary signaling pathways for cells, decoding their coordinated actions through network analyses and pattern detection methods. Using manifold learning and quantitative assessments, CellChat classifies signaling routes and differentiates between universal and context-specific pathways across diverse datasets. Consequently, it aids in revealing novel intercellular communications and in building communication blueprints for a range of tissues (22).

## Materials and methods

### Single-cell and Single-nucleus RNA-seq Data Analysis

#### 1. Data Collection

The scRNA-seq and snRNA-seq data from human liver and Peripheral Blood Mononuclear Cell (PBMC) of NAFLD and healthy controls were collected from the NCBI GEO (until June 2023). Only datasets generated with 10x Genomics Single-Cell Reagent Kit were included.

#### 2. Data Alignment

The FASTQ-formatted data were downloaded and converted into feature, barcode and matrix with the human GRCh38 genome using ‘cellranger’ (cellranger-6.1.2). For snRNA-seq, intron was included for quantification (23). The doublets were removed with the R package ‘DoubletFinder’ (DoubletFinder-2.0.3) for each sample.

#### 3. Data Normalization and Clustering

With the R package ‘Seurat V5’ (Seurat 4.9.9.9042) (https://satijalab.org/seurat/articles/seurat5_integration.html), the datasets were integrated and processed using the anchor-based reciprocal principal components analysis (RPCA) method. After normalization and batch correction, the scRNA-seq and snRNA-seq data underwent unsupervised clustering.

For normalization, the ‘LogNormalize’ method was utilized with a default scale factor of 10,000, and subsequent feature identification and data scaling were done by default. The scaled data was then subjected to Principal Component Analysis (PCA) with a default setting of computing and storing 50 Principal Components (PCs). To perform integration in low-dimensional space, we selected the anchor-based RPCA integration method, which aimed to co-embed shared scRNA-seq and snRNA-seq library types across batches, yielding a dimensional reduction (integrated.rpca).

For cell clustering, a shared nearest neighbor (SNN) modularity optimization-based algorithm was employed, with the dimensions of reduction set to 1:15 and the resolution parameter set to 0.2 (liver) and 0.1 (PBMC).

#### 4. Cell Type Annotation and Data Analysis

We annotated liver and PBMC clusters with canonical markers by manually curated the information to enhance the accuracy of gene annotation. To gain a more nuanced understanding of transcriptomic variations, we further segmented certain cell types in additional supplementary files. This enabled us to identify subtype-specific differences in both healthy and NAFLD conditions. We then compared the expressions of potential NAFLD biomarkers across multiple clustering methodologies and depicted the results on distinct UMAP plots. For subtype cell clustering, a shared nearest neighbor (SNN) modularity optimization-based algorithm was employed, with the dimensions of reduction set to 1:15 and the resolution parameter set to 0.05 (HSC/MFB).

#### 5. Cell Communication Analysis

The sc- and snRNAseq data of NAFLD liver and PBMC were merged together and RPCA integrated before normalization. A Seurat object with a new assay of 5,000 cells in each cell type in a tissue was subsetted. The R package ‘CellChat’ (CellChat 1.6.1) was utilized to analyze the data. We refined the CellChatDB database to include only interactions involving secreted signaling, as the candidate NAFLD biomarkers originate from genes that are secreted.

#### 6. Database Construction

To visualize gene expression variation during the development of NAFLD severity at the cell level, we generated a NAFLD sc- and snRNA-seq database (NAFLD-DB) using the tool ‘ShinyCell’. (https://github.com/SGDDNB/ShinyCell). This database was deployed at https://dreamapp.biomed.au.dk/NAFLD-scRNA-seq/.

### Validation of QSOX1 and IL1RAP as Biomarkers in NAFLD at the Liver Cell Level

#### 1. Study Design

Our objective was to confirm the *QSOX1* and *IL1R A* g*P*ene expression in cells of different severity of NAFLD. To address this question, we performed immunohistochemical staining (IHC) for liver QSOX1/IL1RAP on a group of NAFLD patients from Shenzhen Traditional Chinese Medicine Hospital, China (SZTCMH). Details of this cohort have been provided previously (16).

#### 2. Human Samples and IHC

For IHC, we collected 12 fixed liver tissues from archived histological samples at SZTCMH between 2014 and 2023, and details were in our earlier study (16).

#### 3. Statistical Analysis

The significance for all statistical tests was two-sided, with P < 0.05. All data analysis was presented in the plots using R-4.3.1. Statistical analysis in the Violin plots was performed with Wilcoxon rank sum test to assess differences between unpaired groups.

## Results

### Alignment of ScRNA-seq and snRNA-seq Data

We incorporated twelve datasets (GSE122083, GSE125188, GSE136103, GSE144430, GSE159977, GSE169446, GSE175793, GSE179886, GSE182159, GSE189539, GSE212837 and GSE217235) into our study. The scRNAseq and snRNA-seq data encompass liver and PBMC samples of both healthy controls and patients with NAFLD (**Supplementary Table 1**). The detailed information regarding the data alignment was presented in the **Supplementary Table 2.**

By employing RPCA integration, we achieved precise comparative analyses across single-cell and single-nucleus sequencing datasets. To address batch effects arising from diverse sequencing technologies and studies involving various experimental batches, donors, and conditions, we conducted integration and normalization, as depicted in **Figure 1A**. After filtering out doublets, the dataset was refined, resulting in a total of 427,787 liver cells (including 214,795 healthy cells and 212,992 NAFLD cells) and 122,238 PBMC cells (including 92,319 healthy cells and 29,919 NAFLD cells) available for subsequent analysis.

**Figure 1.**
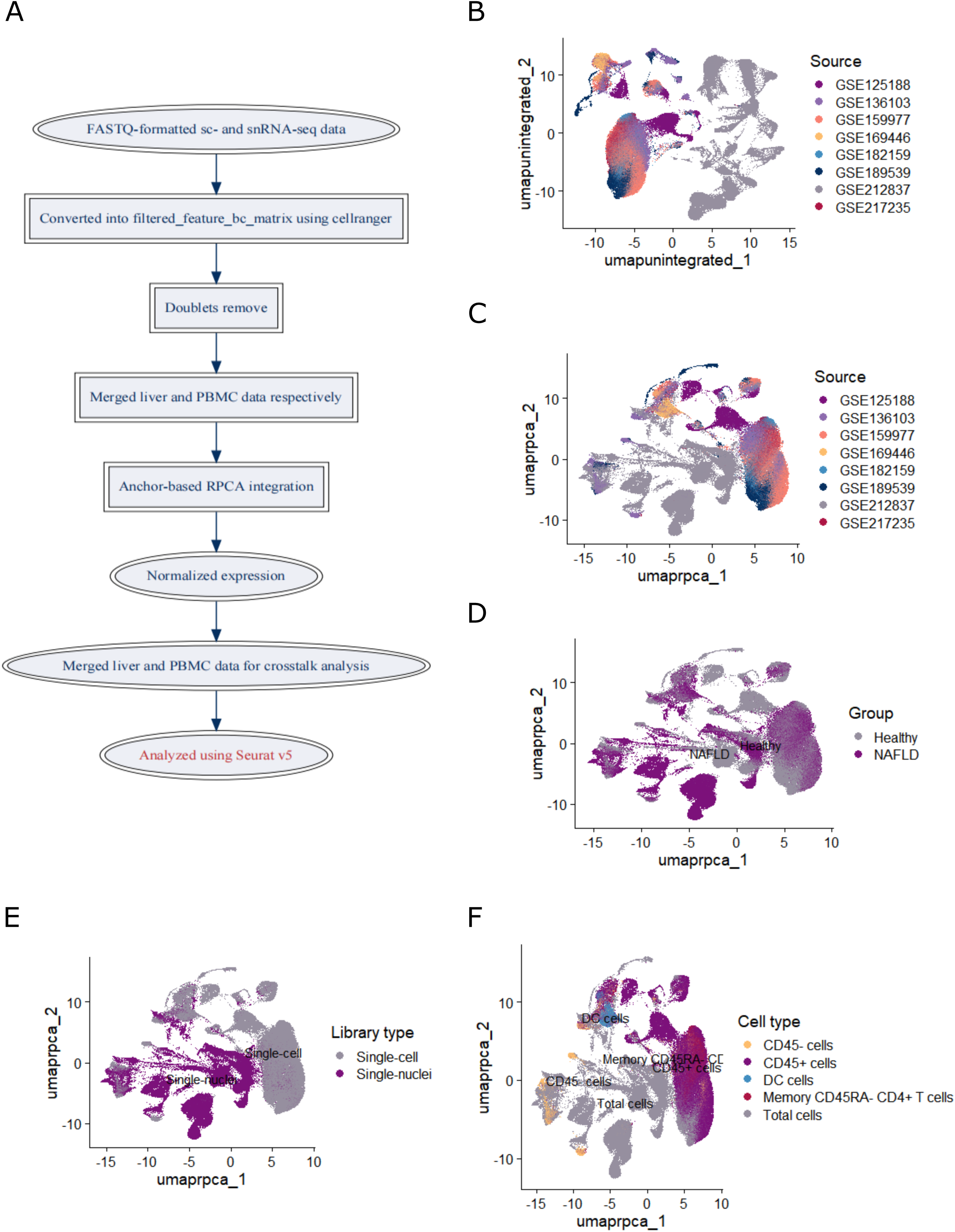
Liver single-cell and nucleus RNA sequencing data integration. A. Illustration of data processing. B. Uniform Manifold Approximation and Projection (UMAP) based on the unintegrated data. C. UMAP based on the anchor-based reciprocal principal components analysis (RPCA) integrated data. D. UMAP based on the health and NAFLD groups after RPCA integration. E. E. UMAP based on the library type after RPCA integration. F. UMAP based on the input cell type after RPCA integration.

### The Effectiveness of Integration and Normalization on Liver Transcriptome Profiles

PCA demonstrated the effectiveness of the RPCA integration and normalization process in eliminating noticeable batch effects of liver transcriptome profiles compared to the unintegrated UMAP, as depicted in **Figure 1B and 1C**. Moreover, sample separation was not primarily driven by health/NAFLD group or library type, as indicated in **Figure 1D and 1E**. Instead, cell type emerged as the primary factor influencing transcriptome profiles, as illustrated in **Figure 1F**.

### Unsupervised Clustering and Annotation of Liver Cells and Nuclei Data

Initially, we categorized all 427,787 liver cells into 12 distinct cell types using gene markers. The identified cell types were annotated using signatures of known canonical markers (**Figure 2A, B; Supplementary Table 3**).

**Figure 2.**
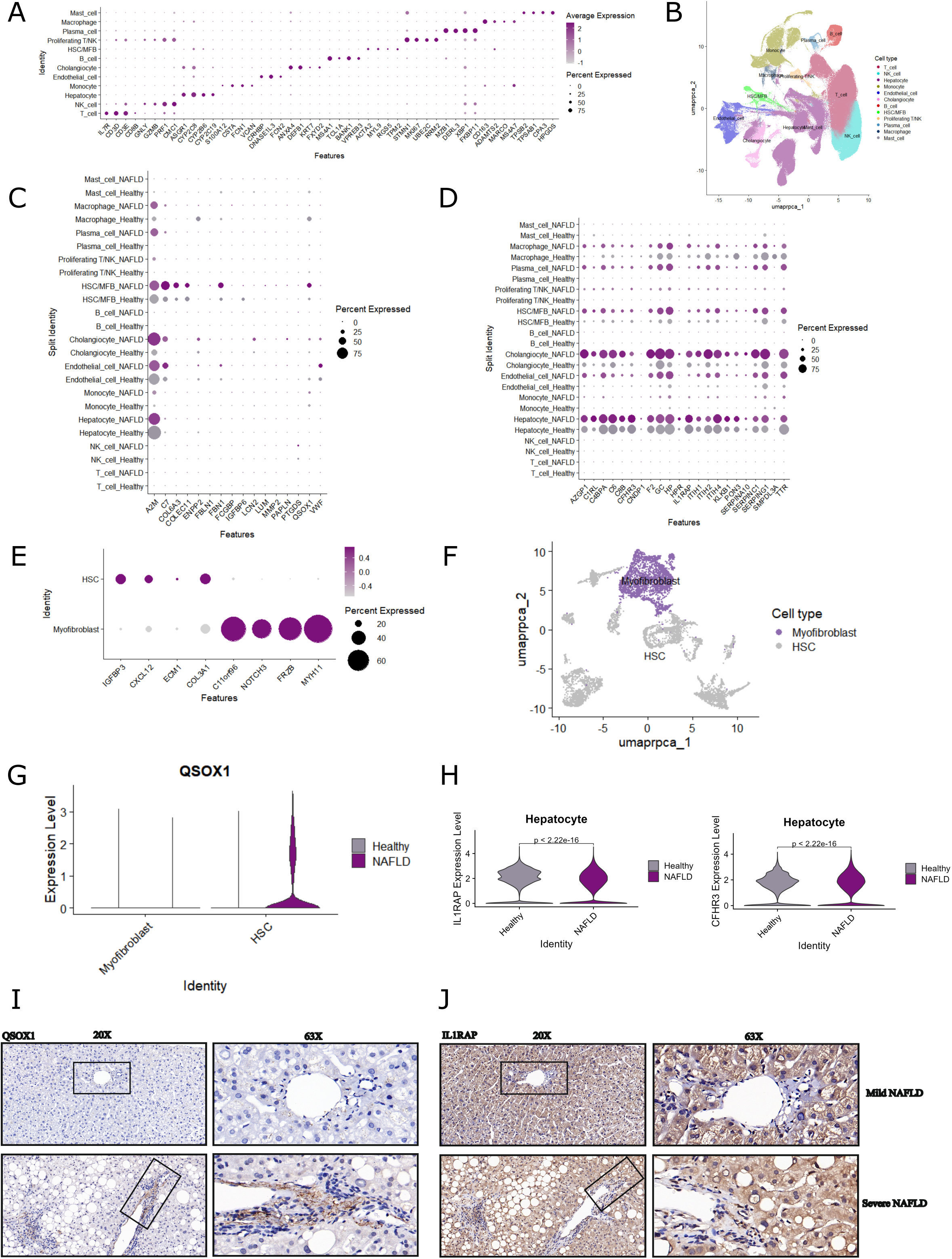
Integrated liver transcriptome data annotation, analysis and verification. A. Gene markers of liver cell types. B. Total 12 distinct cell types were clustered according to the canonical markers. C. Dot plot of 16 up-regulated secreting marker expressions in cell clusters. D. Dot plot of 22 down-regulated secreting marker expressions in cell clusters. E. Gene markers of HSC (hepatic stellate cell)/MFB (myofibroblast) cell types. F. Total 2 distinct cell types were clustered according to the canonical markers. G. Violin plot of QSOX1 gene expressions in the hepatic stellate cell and myofibroblast. H. Violin plot of IL1RAP, CFHR3 gene expressions in the Hepatocytes. I. Representative immunohistochemical staining (IHC) images of QSOX1 in liver biopsies from mild NAFLD patients (N0-4, F0-2) and severe NAFLD patients (N5-8, F3-4) in our previous research (16). J. Representative IHC images of IL1RAP in liver biopsies from mild NAFLD patients (N0-4, F0-2) and severe NAFLD patients (N5-8, F3-4) in our previous research (16). Statistical testing was performed using the Wilcoxon rank sum test, with p-values shown in the plot.

### Tracing the Cellular Origins of Promising Biomarkers for Liver NAFLD Severity

In our previous study (16), we pinpointed key genes that are significantly involved in the advancement of NAFLD. Specifically, we discovered 16 genes, including A2M, C7, COL6A3, COLEC11, ENPP2, FBLN1, FBN1, FCGBP, IGFBP6, LCN2, LUM, MMP2, PAPLN, PTGDS, QSOX1, and VWF, that exhibited elevated secretory activity correlating with the severity of NAFLD. On the other hand, we also identified another set of 22 genes, namely AZGP1, C1RL, C4BPA, C6, C8B, CFHR3, CNDP1, F2, GC, HP, HPR, IL1RAP, ITIH1, ITIH2, ITIH4, KLKB1, PON3, SERPINA10, SERPINC1, SERPING1, SMPDL3A, and TTR, that showed reduced secretory activity, also in relation to the severity of the condition. These particular genes were not only implicated through RNA-seq data but were also corroborated by proteomics datasets. The correlation was such that the expression patterns of these genes seemed to escalate or diminish in tandem with increasing NAFLD severity. We investigated how these genes contribute to NAFLD pathogenesis, and why their particular expression patterns are so closely associated with the progression of NAFLD.

Delving into the intricate cellular milieu, the expression profiles of these potential biomarkers were meticulously examined across diverse liver cell categories. It was observed that the HSC/MFB cellular contingent distinctly exhibited pronounced expression levels of C7, COL6A3, FBN1, and QSOX1 compared to other cellular phenotypes. Intriguingly, these molecular markers were conspicuously prevalent in NAFLD-afflicted cohorts over their healthy counterparts (**Figure 2C**). In light of the overwhelming majority of hepatocytes in hepatic architecture, significant expression of IL1RAP, ITIH2, TTR, and SERPINA10 was ascertained in these cells, spanning all identified subtypes. This expression spectrum was notably skewed towards the healthy clusters versus NAFLD groups, intimating a palpable downregulation at the holistic hepatic expression matrix (**Figure 2D**).

Cutting-edge developments in medical science have clarified the primary source of myofibroblasts, identifying hepatic stellate cells (HSCs) as their main origin (24, 25). Functioning as lipid storage entities, HSCs share considerable similarity with adipocytes, or fat cells, particularly when the liver is functioning optimally (26). This newfound insight broadens our understanding of liver diseases, including liver fibrosis and cirrhosis, where myofibroblasts play a significant role. Interestingly, the elevated levels of QSOX1, which is enriched in the HSC/MFB, correlate with both the NAS score and fibrosis score, offering a potential biomarker for NAFLD severity.

A more detailed exploration of the HSC/MFB clustering, and annotation using canonical markers has revealed two distinct subsets: hepatic stellate cells and myofibroblasts (**Figure 2E, F; Supplementary Table 3**). Within this context, QSOX1 manifested heightened expression within the NAFLD subset of HSC (**Figure 2G**). In contrast, IL1RAP and CFHR3, bearing the torch for the loftiest expression within hepatocytes amongst all subtypes, evinced diminished expression within the NAFLD demographic compared to its healthier cohort (illustrated in **Figure 2H**).

In a supplementary validation, liver biopsies sourced from patients with varying NAFLD severity were subjected to IHC in our antecedent study (16). Conclusive findings reinforced that QSOX1 resonates with heightened expression specifically in HSC/MFB cells of severe NAFLD patients (**Figure 2I**), while IL1RAP’s expression was attenuated in hepatocytes of these patients as seen in **Figure 2J** (16).

### The Effectiveness of Integration and Normalization on PBMC Transcriptome Profiles

To explore the expression of biomarkers in PBMC between NAFLD patients and healthy controls, we tracked their expression variations using RPCA integrated scRNA-seq data. The results from PCA showed that normalization successfully mitigated discernible batch effects compared to the unintegrated data, as shown in **Figure 3A and B**. The primary factors driving sample differentiation were not the health and NAFLD group, as depicted in **Figure 3C**. Instead, the cell type surfaced as the primary determinant impacting transcriptome profiles, as illustrated in **Figure 3D**.

**Figure 3.**
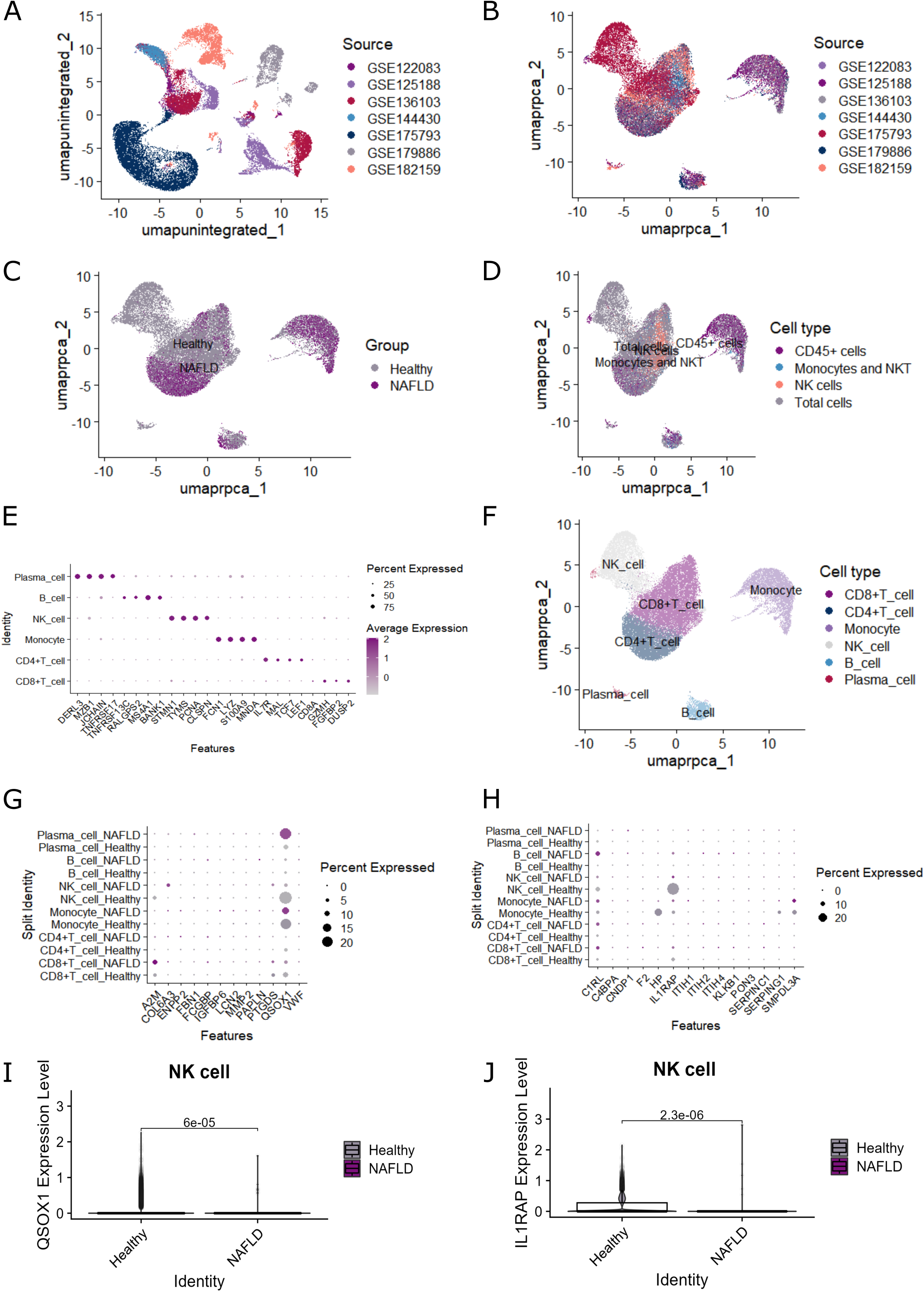
Tracing the Cellular Origins in Liver and PBMC Data Integration. A. Uniform Manifold Approximation and Projection (UMAP) based on the unintegrated data.Dot plot of 16 up-regulated secreting marker expressions in cell clusters. B. UMAP based on the anchor-based reciprocal principal components analysis (RPCA) integrated data. C. UMAP based on the health and NAFLD groups after RPCA integration. D. D. UMAP based on the input cell type after RPCA integration. E. Gene markers of PBMC cell types. F. Total 6 distinct cell types were clustered according to the canonical markers. G. Dot plot of 16 up-regulated secreting marker expressions in cell clusters. H. Dot plot of 22 down-regulated secreting marker expressions in cell clusters. I. Violin plot of QSOX1 gene expressions in the NK cell. J. Violin plot of IL1RAP gene expressions in the NK cell. Statistical testing was performed using the Wilcoxon rank sum test, with p-values shown in the plot.

### Unsupervised Clustering Identifies Clusters of PBMC Cells

We first clustered all the 122,238 PBMC cells into 6 cell types, and the identified cell types were annotated using signatures of known canonical markers (**Figure 3E, F; Supplementary Table 3**).

### Tracing the Cellular Origins of Promising Biomarkers for NAFLD severity in PBMC

We identified 16 genes that are up-regulated (A2M, C7, COL6A3, COLEC11, ENPP2, FBLN1, FBN1, FCGBP, IGFBP6, LCN2, LUM, MMP2, PAPLN, PTGDS, QSOX1, VWF) and 22 that are down-regulated (AZGP1, C1RL, C4BPA, C6, C8B, CFHR3, CNDP1, F2, GC, HP, HPR, IL1RAP, ITIH1, ITIH2, ITIH4, KLKB1, PON3, SERPINA10, SERPINC1, SERPING1, SMPDL3A, TTR) with increasing severity of NAFLD, as evidenced by both RNA-seq and proteomics data. In PBMC, QSOX1 and IL1RAP have the highest expression levels in Natural killer cells (NK cells) across all subtypes. Interestingly, healthy individuals showed higher expression of these genes than those with NAFLD (**Figure 3G, H**). Nonetheless, the baseline expression levels of these genes in PBMC were too faint to support a significant comparison. The low expression within PBMC may not affect their detectability and proportional concentrations in plasma. Even when a gene exhibits varied expression in a distinct cell type or tissue, it’s not a given that there will be discernible differences in broader systems like plasma, especially when the initial expression is inherently low (**Figure 3I, J**).

### The intercellular crosstalk in the liver and PBMC of NAFLD patients and healthy controls revealed by integrated sc- and snRNAseq data analysis

Since our candidate biomarkers were derived from secreted genes, it was essential to delve deeper into the communication mechanisms of these genes. We utilized the ‘CellChat’ R package to analyze the ligand-receptor interactions, enabling us to better understand the crosstalk of secreted signaling and their potential implications in the biological processes of NAFLD.

The integrated RPCA analysis combined a total of 427,787 liver cells and 122,238 peripheral blood mononuclear cells (PBMCs). The data was then partitioned into subsets comprising 5,000 cells for each cell type within the respective tissues. Subsequent analysis was carried out using the ‘CellChat’ R package (**Figure 4A**). The examination of ligand-receptor pairs of secreted signaling, which constitutes 61.8% of all categories as depicted in **Figure 4B**, within the transcriptomic data from the liver and PBMCs in the NAFLD group indicates a strong correlation. Specifically, the interaction between IL1B-(IL1R1+IL1RAP) in liver monocytes and hepatocyte/cholangiocyte may provide a convincing rationale for the correlation between NAFLD severity and the down-regulated expression of IL1RAP in the liver, as highlighted in **Figure 4C and F**. In our earlier research (16), we pinpointed potential diagnostic markers from candidate genes by juxtaposing both up-regulated and down-regulated gene clusters against the secretome database from the Human Protein Atlas. Through this cross analysis, we discovered 349 genes that encode secreted proteins: 249 of these genes were up-regulated, while 100 were down-regulated. Of the totally 349 secreted genes, both FGFR2 and JAM3 exhibited up-regulation, while ERBB3 and SDC4 were notably down-regulated. These four genes are cataloged within the ligand-receptor pairs in the CellChat database, as referenced in **Supplementary Table 4**. Yet, when examining the crosstalk analysis derived from the sc- and snRNAseq data of NAFLD patients, only the down-regulated SDC4 (ANGPTL4-SDC4) and ERBB3 (NRG1-ERBB3) emerged as significant receptors in these pairs (see **Supplementary Tables 4 and 5**). The ANGPTL4-SDC4 pairing occurs between liver hepatocytes and hepatocyte/cholangiocyte, as depicted in **Figures 4D and 4G**. Similarly, the NRG1-ERBB3 interaction is evidenced between liver hepatocytes and cholangiocytes, highlighted in **Figures 4E and 4G**. An accompanying violin plot reveals expression patterns of the three crucial down-regulated secreted genes from the crosstalk analysis. Intriguingly, Most of them are predominantly found in hepatocytes, as demonstrated in **Figure 4H**. It may intricate that the significant down regulation of liver gene expression in bulk RNA sequencing may mostly caused by the gene variation in hepatocyts since it is the biggest population in the liver. And the down-regulation may caused by the ligands acted on the receptors in the hepatocytes when the NAFLD severity developed.

**Figure 4.**
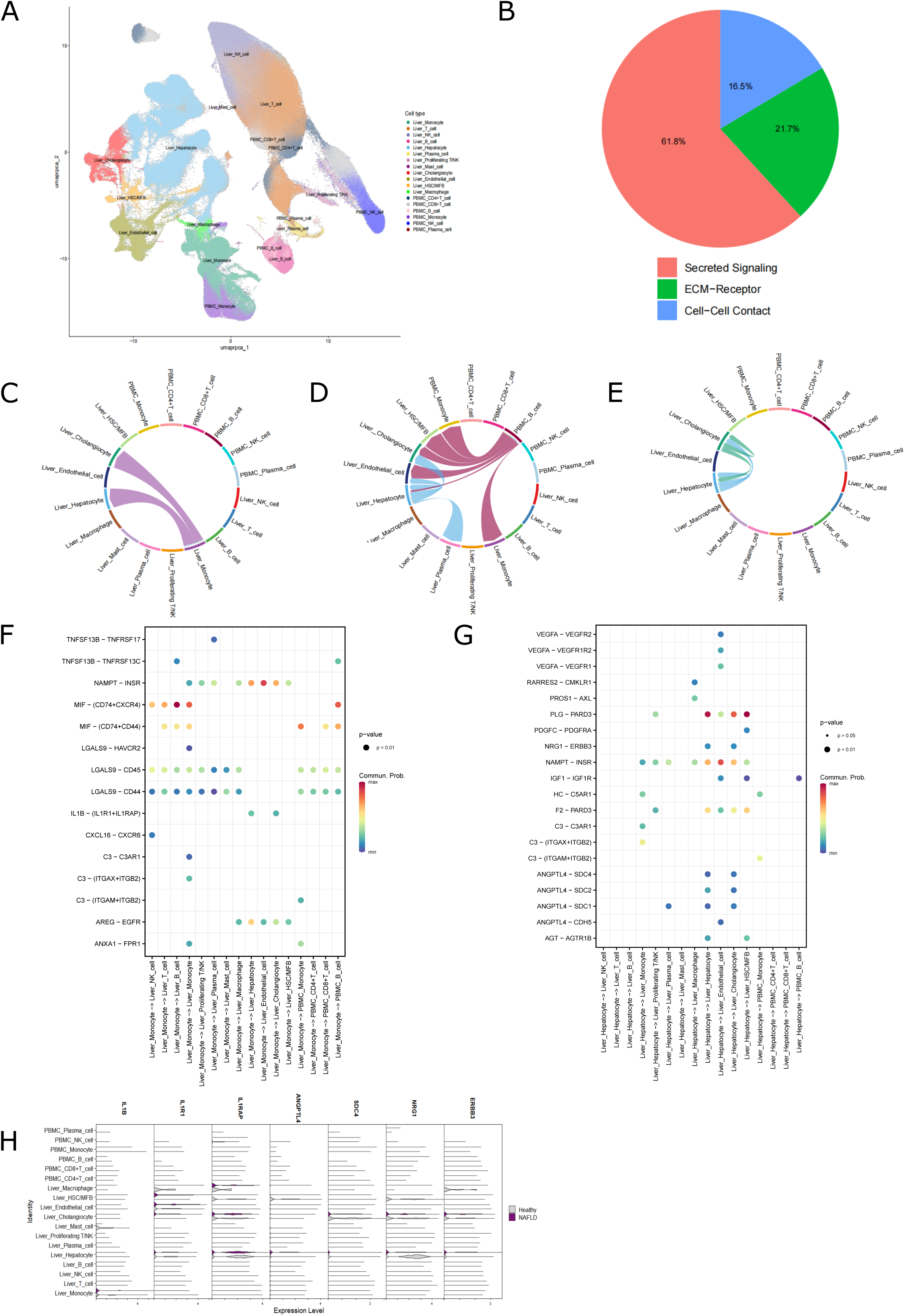
The cell communication in the liver and PBMC transcriptomics data. A. UMAP based on the anchor-based reciprocal principal components analysis (RPCA) integrated liver and PBMC data. B. Ligand-receptor pairs of secreted signaling, which constitutes 61.8% of all categories. C. The interaction between IL1B-(IL1R1+IL1RAP) in liver monocytes and hepatocyte/cholangiocyte. D. The ANGPTL4-SDC4 pairing occurs between liver hepatocytes and hepatocyte/cholangiocyte. E. The NRG1-ERBB3 interaction is evidenced between liver hepatocytes and cholangiocytes F. The interaction between IL1B-(IL1R1+IL1RAP) in liver monocytes and hepatocyte/cholangiocyte. G. The ANGPTL4-SDC4 pairing occurs between liver hepatocytes and hepatocyte/cholangiocyte. H. Violin plot of IL1B, IL1R1, IL1RAP, ANGPTL4, SDC4, NRG1, ERBB3 gene expressions in the liver and PBMC.

## Discussion

The inherent complexities of NAFLD pathogenesis consistently propel scientists to extend the boundaries of traditional diagnostic and investigative methodologies, thus underscoring the escalating importance of genomics technology within this domain. Our research RPCA integrated publicly available scRNA-seq and snRNA-seq datasets to identify potential biomarkers, thereby contributing to the enhanced understanding of the intricate pathogenic processes driving NAFLD (27). Using this approach, we corroborated the significant correlation between the expression ratio of QSOX1/IL1RAP and the severity of NAFLD at the cellular level (16). Furthermore, through an analysis of the integrated large-scale scRNA and snRNA sequencing data to interpret potential biomarkers, we shed light on previously undiscovered facets of NAFLD pathogenesis and intercellular crosstalk.

QSOX1 and its less common variant, QSOX2, are unique among sulfhydryl oxidases because they are specifically found in the Golgi apparatus of animal cells (28). Studies found elevated expression of QSOX1 in quiescent fibroblasts, and it is also released into the bloodstream (29, 30). QSOX1 plays a crucial role in the proper construction of the ECM, particularly elements similar to the basement membrane (BM) in cell culture. The BM, an ECM layer interfacing bodily cavities or blood vessels with underlying stromal fibroblasts, is largely composed of elements generated by fibroblasts (31). The exact reason behind QSOX1’s secretion— whether due to overwhelming the Golgi’s retention mechanism or some specialized secretion pathway—remains unclear. Our findings highlight an increased expression of QSOX1 in fibroblasts within NAFLD-affected liver tissue, suggesting that Golgi function in fibroblasts could be a relevant focus for NAFLD research (32).

Fibrogenesis is a key driver in NAFLD progression and is chiefly managed by specialized cells called myofibroblasts (33, 34). While these cells are not present in a healthy liver, they can quickly develop following liver damage. Emerging research has established that these myofibroblasts predominantly originate from hepatic stellate cells (HSCs) (24, 25), which act mainly as lipid storage cells and bear resemblance to fat cells, or adipocytes, under healthy conditions (26).

In our previous article (16), the GO analysis revealed significant enrichment of the up-regulated genes involved in the fibrosis-related process, such as ECM organization. Similar to QSOX1 in terms of expression pattern and location, COL6A3 is associated with liver fibrosis and also plays a role in the ECM organization (35). Increased production of ECM moieties by diverse liver cell types is tightly linked with hepatic fibrosis. Research has demonstrated that adipocytes are affected by the mechanics of their surrounding ECM, such as stiffness and architecture, independent of initial ligand density. In response to a stiffened, modified ECM structure, adipocytes enhance the expression of fibrotic genes and increase ECM deposition. This reaction eventually results in a dysfunctional, fibrotic state (36, 37). This represents a significant research trajectory in the ongoing investigation of NAFLD pathophysiology.

IL1RAP is a crucial co-receptor expressed in numerous tissues and cell types, emphasizing its pivotal role in immune responses, cancers, and infectious diseases (38). The IL1RAP serves as a subunit to the type I IL-1 receptor (IL-1RI), a specific plasma membrane receptor, which facilitates the transmission of numerous biological effects of the pro-inflammatory cytokine IL-1 on an array of target cells (39, 40). Research indicates that one of the isoforms of IL1RAP, known as the soluble isoform of the IL-1 receptor accessory protein (sIL1RAP), has been found to have lower levels in the plasma of obese individuals when compared to their non-obese subjects (17). IL1RAP, enriched in the cytoplasm of liver cells and released into the bloodstream, shows variations in expression levels that correlate with the severity of NAFLD, as per our study. Nevertheless, its specific role in NAFLD progression remains an enigma.

CFHR3, displaying similar expression patterns and localization as IL1RAP, also warrants further investigation for its potential role in diagnosing NAFLD (30). The complementary DNAs (cDNAs) that encode CFHR3 were extracted from a human liver cDNA library. These have been located within the ‘regulators of complement activation’ (RCA) gene cluster situated on human chromosome 1 (41, 42). CFHR3 has been linked with some diseases, including hepatocellular carcinoma (HCC), although the underlying mechanism is still not clearly understood (43). Our research indicates that CFHR3 expression decreases in correlation with the severity of NAFLD, including both the NAS and fibrosis scores. This finding opens a new avenue of research into the pathophysiology of NAFLD.

The IL1 pathway, especially IL1B, has attracted significant attention and has been extensively researched within the realm of NAFLD (35, 44–47). IL-1β exacerbates liver steatosis, inflammation, and fibrosis by interacting with the IL-1 receptor, which is extensively expressed across various liver cell types (44). In 2021, Unamuno et al. discovered that in the liver of obese patients with NASH, there was a significant upregulation of mRNA levels for NLRP3 (PL<L0.05) and its associated proinflammatory mediator IL1B (PL<L0.01) (35). In 2018, Pan and colleagues showed that in NASH patients, IL-1β is primarily secreted by Kupffer cells, which are a type of monocyte in the liver. This secretion is driven by the activation of the NLRP3 inflammasome, stimulated by mitochondrial DNA (48). From our studies on cellular interactions within NAFLD, we have formed and to some extent validated a hypothesis that the IL-1β, when secreted, serves as a ligand affecting the receptors IL1R1 and IL1RAP in both hepatocytes and cholangiocytes. This interaction between the ligand and its receptors seems to play a crucial role in modulating the expression of IL1RAP, which is closely associated with NAFLD severity. The dynamic interplay between these cellular components provides a deeper understanding of the disease’s underlying mechanisms and offers potential avenues for NAFLD biomarkers and therapeutic interventions.

The utility of Seurat v5 in processing and integrating scRNA-seq and snRNA-seq data is apparent in our study. However, we must also acknowledge the inherent limitations of these techniques (49). Therefore, our findings must be interpreted with caution, and further confirmatory studies are required to validate the candidate biomarkers and the NAFLD pathogenesis.

Utilizing the ‘CellChat’ R package allowed for a streamlined analysis of the data. Given the vastness of the data, it was segmented into more digestible portions, sketching them into clusters of 5,000 cells each, ensuring a more organized scrutiny. An outstanding revelation from this analysis was the significant presence of ligand-receptor pairs of secreted signaling within the NAFLD group, which constituted a remarkable 61.8% of all categories. This prevalence underscored their crucial role in the interactions between liver and PBMC cells. The interaction of IL1B-(IL1R1+IL1RAP) between liver monocytes and hepatocyte/cholangiocyte was of particular interest, suggesting a plausible explanation for the detected decline in IL1RAP expression in the liver - a potential indicator of NAFLD severity. Referring back to our prior research, the crosstalk analysis, built on sc- and snRNAseq data from NAFLD patients, spotlighted SDC4 (in association with ANGPTL4) and ERBB3 (aligned with NRG1) as receptors of importance. The relationships between ANGPTL4-SDC4 and NRG1-ERBB3 with liver hepatocytes and hepatocyte/cholangiocytes, respectively, reflect their likely significance in the liver’s cellular dynamics. This proposition is bolstered by a violin plot that vividly illustrates the expression patterns of three crucial down-regulated secreted genes from the crosstalk analysis, with hepatocytes prominently featuring as their central hub.

Syndecan-4 (SDC4) is a transmembrane protein that is glycosylated and prevalent in numerous tissues (50). Essential cellular functions such as differentiation, adhesion, migration, and prognosis are facilitated by SDC4, given its ability to bind to a variety of external molecules and engage in signal transmission (51–53).

ERBB3, or Receptor tyrosine-protein kinase erbB-3, is a membrane-associated protein from the EGFR/ERBB family of receptor tyrosine kinases. Research has shown its involvement in numerous liver conditions, including liver fibrosis (54, 55).

In conclusion, our research sheds light on the intricacies of NAFLD pathogenesis and highlights the significance of newly identified biomarkers in tracking disease progression. These insights serve as a foundation for subsequent studies delving into the molecular intricacies of NAFLD, all aiming towards enhancing disease diagnosis, management, and the outcomes of patients.

## Authors’ contributions

WFM and LL designed the study and interpreted the data. The analysis strategy has been developed by LL and WFM. WFM and LL collected, analyzed, and interpreted the data. WFM drafted the manuscript. YLL, HG, and MBK revised the manuscript. WFM and LL performed data analysis and interpretation. BQC, JRH, MLL, XZ, SMX, BLZ, QL, QH, MQM contributed to the validation of the candidate biomarkers. Technical support: LL and JRH. All authors reviewed and approved the final version of the manuscript.

## Data and code availability statement

The data and code that support the findings of this study are available at https://github.com/cynthia139/NAFLD-scRNA-seq.

## Data Availability

https://dreamapp.biomed.au.dk/NAFLD-scRNA-seq/

## Abbreviations

avg_log2FC: log fold-change of the average expression between the two groups
BM: basement membrane
cDC1: type 1 conventional dendritic cell
cDNAs: complementary DNAs
ECM: extracellular matrix
F: Fibrosis score
GEO: Gene Expression Omnibus
GO: Gene Ontology
GRCh37: Genome Reference Consortium Human Build 37
HCC: hepatocellular carcinoma
HSCs: hepatic stellate cells
IHC: immunohistochemistry staining
IL-1RI: type I IL-1 receptor
IL1RAP: Interleukin-1 receptor accessory protein
LECs: lymphatic endothelial cells
N: NAS score
NAFL: Non-alcoholic Fatty Liver
NAFLD: Non-alcoholic fatty liver disease
NAFLD-DB: NAFLD gene expression database
NAS: NAFLD activity scores
NASH: Non-alcoholic Steatohepatitis
NK cells: Natural killer cells
PBMC: Peripheral blood mononuclear cell
PCA: Principal components analysis
PCs: Principal Components
QSOX1: Quiescin sulfhydryl oxidase 1
RCA: regulators of complement activation
RNA-seq: RNA sequencing
RPCA: reciprocal principal components analysis
scRNA-seq: single-cell RNA sequencing
sIL1RAP: the soluble isoform of the IL-1 receptor accessory protein
SNN: shared nearest neighbor
snRNA-seq: Single-nucleus RNA sequencing
SZTCMH: Shenzhen Traditional Chinese Medicine Hospital, China
UMAP: Uniform Manifold Approximation and Projection.

